# Impact of intestinal dysbiosis on clinical course in severe acute pancreatitis: a multicenter prospective observational study

**DOI:** 10.1101/2023.03.30.23287938

**Authors:** Takehiko Oami, Shigenobu Ishida, Taku Oshima, Akinobu Yamamoto, Taka-aki Nakada

## Abstract

**Background:** Dysbiosis, an imbalance of the intestinal microbiota in critically ill patients, reportedly contributes to poor outcomes. Controlling the disruption of intestinal homeostasis could be promising tactics for infectious complications in acute pancreatitis. To improve the mortality rate of severe acute pancreatitis with high frequency of infectious complications, it is warranted to elucidate the pathophysiology for development of new treatment strategies. We hypothesized that patients with severe acute pancreatitis would demonstrate gut dysbiosis, leading to an onset of infectious complications and poor outcomes.

**Methods:** We will conduct a prospective study to compare the sequential changes in intestinal microbiota using 16s RNA metagenomics and metabolomics between patients with severe acute pancreatitis and mild acute pancreatitis. We will enroll adult patients (18 years of age or older) diagnosed with acute pancreatitis and newly admitted to the hospitals for 48 hours or longer. We will exclude patients with inflammatory bowel disease, patients with diarrhea prior to admission, patients who have received antimicrobial agents for more than 1 week in the 2 months prior to admission. We will collect stool and blood samples on day 1 and 6. The primary outcome is changes in various parameters of the intestinal microbiota, protein concentration in stool, and metabolite concentration. The secondary outcomes include relationship between each parameter and short- and long-term prognosis, correlation of each parameter with treatment details and clinical course during ICU stay, and associations among the amount of diarrhea and alpha-diversity parameters, protein concentration in each stool, and metabolite concentration.

## Background

Severe acute pancreatitis is one of the leading causes of multiple organ failure due to spread of inflammation to adjacent organs and systemic inflammatory response syndrome (SIRS) [1]. As the efficacy of specific medication has not been proven, intensive care, including initial infusion therapy, control of secondary infections, and artificial support therapy, is the mainstay of treatment. The mortality rate of severe acute pancreatitis in Japan is as high as approximately 10%, and the rate of late mortality after 2 weeks of illness is high [2]. Infectious complications account for a large proportion of late mortality. To improve the life-saving rate of severe acute pancreatitis, it is necessary to elucidate the pathogenesis of infectious complications and to develop new treatment methods.

Metagenomics using next-generation sequencers has revealed intestinal dysbiosis, an imbalance of the microbiota triggered by infection or administration of antimicrobial agents, occur during clinical course of acute pancreatitis [3]. Infectious complications, such as infectious pancreatic necrosis, have been reported to contribute to poor outcomes and longer hospital stay [4]. Therefore, it is reasonable to investigate the relationship between an alteration of gut microbiota and an onset of infections complications during the clinical course of severe acute pancreatitis.

A study analyzing stool samples from patients with severe acute pancreatitis reported that the bacterial flora presented a decrease in α-diversity and changed composition (β-diversity), which was associated with the severity of pancreatitis [3]. In addition, the intestinal mucosal damage was exacerbated by the increased degradation of short-chain fatty acids such as butyrate due to the progression of intestinal dysbiosis [5]. Recent studies have reported a decrease in the complication rate of infection in severely ill patients treated with probiotics and a favorable outcome after fecal transplantation in severely ill patients who had difficulty controlling infection [6, 7]. To improve the infectious complications of severe acute pancreatitis by controlling the disruption of intestinal homeostasis, including intestinal dysbiosis, without causing adverse events, it is necessary to identify in detail the changes in metabolites produced by intestinal epithelial cells, immune cells, and intestinal bacteria in addition to the characteristic bacterial changes in intestinal dysbiosis.

Therefore, we hypothesized that patients with significant changes in the intestinal microbiota, metabolomes, and intestinal proteins are associated with an onset of infectious complications and mortality in patients with severe acute pancreatitis. The aim of this study is to determine the sequential changes in intestinal microbiota using 16s RNA metagenomics and metabolomics in patients with in severe acute pancreatitis.

## Methods

### Study design and settings

We will conduct a multicenter prospective study to investigate the sequential changes in intestinal microbiota using 16s RNA metagenomics and metabolomics between patients with in severe acute pancreatitis. University hospitals and tertiary care hospitals will participate this study. Inclusion criteria are as follows: Adults (18 years of age or older), patients diagnosed with acute pancreatitis using the Ministry of Health, Labour and Welfare’s criteria for acute pancreatitis severity, patients newly admitted to the participating institution and expected to require hospitalization for 48 hours or longer. We excluded patients with inflammatory bowel disease or diarrhea prior to admission, and patients who have received antimicrobial agents for more than 1 week in the 2 months prior to admission. We will divide enrolled patients into group of severe and mild acute pancreatitis.

This study was approved by the Institutional Review Board of Chiba University Graduate School of Medicine (approval number: M10499) and conducted in accordance with the Ethical Guidelines for Medical and Health Research Involving Human Subjects in Japan. We submitted the review protocol to the pre-print server (medRxiv) and registered with the University hospital medical information network clinical trial registry (UMIN-CTR) [UMIN000050720].

### Sample collection

We will collect stool and blood samples on day 1 and 6. Specimens will be placed in designated containers and stored frozen until further analysis. If it is difficult to collect stool samples, specimens should be submitted by rectal swab. This method has been previously reported and validated in our laboratory. For blood specimens, residual serum from blood tests used in normal practice will be collected. The number of enrolled samples will be 15 patients annually in each institution. We did not conduct a sample size calculation due to the nature of an exploratory study.

### Data collection

We will collect the following patient data: 1) basic patient information: age, sex, height, weight, temperature, 28-day survival, 90-day survival, outcome at discharge, 2) disease information: medical history, diagnosis, treatment details, 3) blood examination results: biochemical tests, blood/electrolytes, coagulation/fibrinolysis, blood gas analysis, blood Interleukin-6 concentration, 4) infection information: the site of infection, causative organism, antimicrobial agents, duration of antimicrobial use, 5) imaging findings: computed tomography, magnetic resonance imaging, ultrasonography, plain radiographs, 6) severity score: Acute Physiologic Assessment and Chronic Health Evaluation (APACHEII), Sequential Organ Failure Assessment (SOFA), 7) nutritional management: enteral feeding, type of enteral feeding, and probiotics administration.

### 16s RNA metagenomics

Deoxyribonucleic acid (DNA) libraries will be created by polymerase chain reaction (PCR) amplification of the V3-4 region of the 16S rRNA gene based on DNA extracted from stool specimens, and the intestinal microbiota is sequenced using a sequencer owned by the RIKEN Center for Biomedical Research. The obtained sequencing data will be analyzed by Qiime2, a bacterial flora analysis software, and finally diversity analysis, Firmicutes/Bacteroidetes ratio, and composition ratio of intestinal flora at the phylum and genus level will be calculated. We will use Linear discriminant analysis E□ect Size (LefSe) determined significant features among groups on Galaxy.

### Predictive functional profiling of microbiota

Using Phylogenetic Investigation of Communities by Reconstruction of Unobserved States (PICRUSt2), we will determine the metabolic functional profiling. Metagenomic Profiles (STAMP) software will be used to analyze predictive functional profiling data.

### Metabolomics

Metabolome analysis will be performed by Liquid Chromatography-Mass Spectrometry (LC-MS) using a mass spectrometer at the Chiba University Shared Instrument Center, focusing on short-chain fatty acids and bile acids after appropriate sample processing.

### Protein assays in stool

We will measure the concentration of calprotectin, IL-6, mucin, and IgA in using frozen stool. Prior to the measurement, the samples will be weighed. The weight of the samples will be used for normalization. We will dissolve the sample in the responsible solutions. The measurement will be performed by the enzyme-linked immunosorbent assay (ELISA).

### Statistical analysis

We will compare the following outcomes between the two groups, patients with mild and severe acute pancreatitis. The primary outcome is changes in various parameters of the intestinal microbiota, protein concentration in stool, and metabolite concentration between patients with severe acute pancreatitis and control patients. The secondary outcomes include relationship between each parameter and short- and long-term outcomes, correlation of each parameter with therapeutic interventions and clinical course during ICU stay, and associations among the amount of metagenomic parameters, protein concentrations in stool, and metabolite concentrations.

The continuous variables are means (standard deviation) or medians (interquartile ranges [IQR]), as appropriate. Student’s *t*-test or Mann–Whitney *U* test will be used according to the normality of distribution. One-way analysis of variance was performed to estimate differences among more than two groups. A *p*-value less than 0.05 is considered to be statistically significant. GraphPad Prism 9 (GraphPad Software, San Diego, CA, USA) will be used for the statistical analysis.

## Data Availability

All data produced in the present study are available upon reasonable request to the authors。

